# Prevalence and associated factors of suicidal behavior among adolescents in rural Bangladesh

**DOI:** 10.1101/2024.07.15.24310423

**Authors:** Rifa Tamanna Mumu, Md Parvez Shaikh, Dipak Kumar Mitra

## Abstract

**Background:** Among the four leading causes of worldwide death, suicide is one, which is prevailing especially in lower-middle-income countries. However, the number of studies is minimal based on adolescent suicidal behavior in rural Bangladesh. This study aims to identify the prevalence and associated factors of suicidal behavior in adolescents in a remote subdistrict in Bangladesh.

**Method:** A cross-sectional study was performed in Lohagara, a rural subdistrict in southern Bangladesh from July, 15 to August 14, 2024. 350 subjects were recruited for the study, all of whom were school-going adolescents aged 11 to 17. The Bengali-translated versions of the Suicidal Behavior-Revised Questionnaire (SBQ-R) and the Depression, Anxiety, and Stress Scale - 21 Items (DASS-21), as well as another structured questionnaire, were used to obtain data from participants. Data were analyzed by STATA version 17.

**Result:** The prevalence of suicidal behavior among adolescents is 12.3% (95% CI: 9.2% to 16.2%) (overall SBQ-R score of 7 or more). Having step-parents, peer conflict, stress, anxiety, and a family history of suicide are found to be significantly associated with the development of suicidal behavior in adolescents. Unmarried and single relationship status and the overprotective attitude of parents play a protective role in growing young adults’ suicidal tendencies.

**Conclusion:** The notable prevalence of suicidal behavior in adolescents underscores the need for screening and intervention at an early age. Different health promotional and educational programs can be organized in schools and communities. The ultimate goal is to protect adolescents with adequate counseling, care, and support.

## 1. Introduction

Approximately, 703 individuals are committing suicide all over the world per year, according to the World Health Organization. Notably, 90% of the world’s adolescents reside in lower-middle-income countries like Bangladesh, where the rate of adolescent suicide is 88%.

Suicide ranks as the fourth predominant cause of death among the populace aged 15 to 29 years. A 9.5% global suicide rate can be observed in males and 4.18% in females among individuals of 15 to 19 years in the year 2009, as well as a greater susceptibility is noticed in elderly adolescents, particularly males (2).

A study carried out with preadults of 12 to 17 years residing in 82 lower-middle-income countries unveiled a 14% rate of suicidal ideation. The percentage was greater in African countries (21%) than in Asia (8%). Factors including female gender, comparatively older age, low socio-economic status, inadequacy of close friendships, overprotective parenting style, peer conflicts, isolation, and victimization, were substantially associated with suicidal ideation and anxiety among young adults (3). In Brazil, a similar trend was discovered, where 11 to 15-year-old preadults manifested a 14.1% prevalence of suicidal ideation, and conducive factors included female gender, conduct disorders, substance abuse, and depressive symptoms (4).

From the perspective of Bangladesh, city dwellers reveal a comparatively declining suicide rate than countryside people. Preadults, the elderly, and rural residents are more prone to suicide, and a 20.1% suicide rate is observed among rural Bangladeshi adolescents (5). In a study conducted with 14 to 19-year-old country adolescents in Bangladesh in 2013, a 5% lifetime suicidal ideation was detected (6). The prevalence of suicidal ideation among university students was 13.8% and influencing factors were gender, year of education, socioeconomic condition, family suicidal history, depression, and observance of traumatic incidents (7). The evaluated percentage of suicidal ideation among 18 to 28-year-olds was 12.8% during the early-COVID-19 period, where the associated factors encompassed prior suicidal attempts, family history, sleep issues, stress, anxiety, and depression (8).

school-going young adults aged 11 to 18 years revealed an 11.7% prevalence of suicidal behavior in the research performed with the Global School-based Health Survey data in Bangladesh (2014). Associated factors included bullying, loneliness, an insufficiency of close friendships, substance abuse, anxiety, experiencing negligence in parental homework checking, participating in sexual activity, and insufficient peer support (9). Although several studies in Bangladesh were conducted to evaluate suicidal ideation in preadults, the concentration was on university students to the greatest extent, which left a gap in recognizing the status among school-going country adolescents. Hence, this study focuses on evaluating the prevalence and influencing factors of suicidal behavior in 11 to 17-year-school-going adolescents in a rural sub-district in Bangladesh. The notion of the current prevalence and associated factors will help stakeholders and policymakers make proper decisions and appropriate actions to build consciousness to safeguard preadults’ mental well-being and protect them from making extreme decisions.

## 2. Method

### 2.1 Study Design and Participants

A cross-sectional study was conducted between July 15 and August 14, 2024, among 11 to 17-year-old adolescents studying in Lohagara Government Pilot High School, a rural high school in Bangladesh. Considering the prevalence of suicidal ideation among adolescents in Bangladesh at 11.7% (9), in 95% confidence interval, and a 4% degree of precision, 350 participants were selected using a stratified sampling technique.

### 2.2 Data Collection Tools

The persistence of suicidal behavior was determined by the Suicidal Behavior-Revised Questionnaire (SBQ-R). A cut-off score of 7 or higher indicated positive screening for having suicidal behavior. The Depression, Anxiety, and Stress Scale-21 Items (DASS-21) was used to assess psychological factors like the level of anxiety, depression, and stress. Another structured questionnaire was used to collect data on participants’ sociodemographic, behavioral, socio-environmental, institutional, and family-environmental factors. All questionnaires were translated into Bengali for effective communication.

### 2.3 Data Management & Analysis

The data for the study was analyzed by STATA version 17. An Independent sample t-test was performed to compare the mean between the two groups and Pearson’s chi-square test was applied to find out the possible association between variables. To adjust the confounding factors, a multivariate analysis using multinomial logistic regression was conducted. Adjusted and unadjusted odds ratios and their 95% CI were used as indicators strength of the association.

### 2.4 Ethical Considerations

Ethical permission was taken from the Institutional Ethics Committee of North South University before data collection (2024/OR-NSU/IRB/0201). A permission letter was obtained from the school authorities. Informed written consent was taken from the legal guardians of students before data collection. The respondents were assured of the confidentiality of information.

## 3. Result

### 3.1 Prevalence of suicidal behavior among adolescents

The Suicidal Behavior-Revised Questionnaire (SBQ-R) was used to evaluate the prevalence of suicidal behavior. An overall cut-off score of 7 or more was used to determine suicidal behavior among adolescents. The mean SBQ-R score was 4.7 (95% CI: 4.3-5). 12.3% (95% CI: 9.2% to 16.2%) (42) of students were found to have suicidal behavior after analysis.***Table 1***.

**Table 1.** Characteristics of 11 to 17-year-old adolescents in Lohagara (a rural sub-district in Bangladesh), 2024.

| Variables | Category | Number<br>(n) | Percentage<br>(%) |
| --- | --- | --- | --- |
| **Sociodemographic** |  |  |  |
| Age | 11-13 years | 115 | 33.7 |
|  | 13-17 years | 226 | 66.3 |
| Gender | Male | 164 | 48.4 |
|  | Female | 175 | 51.6 |
| Occupational involvement besides the study | Involved | 13 | 4.1 |
|  | Not involved | 302 | 95.9 |
| Religion | Hindu | 50 | 14.7 |
|  | Muslim | 290 | 85.3 |
|  | Others | 0 | 0 |
| Number of family members | 4 or less | 173 | 50.7 |
|  | More than 4 | 168 | 49.3 |
| Socioeconomic status | Low | 94 | 27.5 |
|  | Medium | 166 | 48.7 |
|  | High | 81 | 23.8 |
| House ownership of parents | Yes | 292 | 87.6 |
|  | No | 41 | 12.3 |
| Monthly family income (BDT) | >20,000 | 166 | 49.9 |
|  | 10,000-20,000 | 102 | 30.6 |
|  | <10,000 | 65 | 19.5 |
| Living with parents | Both | 266 | 78.7 |
|  | Single parent | 59 | 17.5 |
|  | Without parents | 13 | 3.9 |
| Relationship status | Unmarried and single | 309 | 92.2 |
|  | In a relationship | 15 | 4.5 |
|  | Complex | 9 | 2.7 |
|  | Married | 2 | 0.6 |
| Step-parents | Have | 27 | 8 |
|  | Do not have | 311 | 92 |
| **Behavioral** |  |  |  |
| Smoking | Yes | 7 | 2.1 |
|  | No | 331 | 97.9 |
| Internet use | <3 hours | 321 | 95.3 |
|  | 3-16 hours | 12 | 3.5 |
|  | >16 hours | 4 | 1.2 |
| Extracurricular activities | Involved | 165 | 49.3 |
|  | Not involved | 170 | 50.7 |

| Institutional |  |  |  |  |
| --- | --- | --- | --- | --- |
| Close friends | Have |  | 297 | 87.6 |
|  | Don't have |  | 42 | 12.4 |
| Educational stress | No stress |  | 38 | 11.2 |
|  | Moderate |  | 259 | 76.2 |
|  | Severe |  | 43 | 12.6 |
| Peer conflict | No |  | 242 | 71.6 |
|  | Moderate |  | 88 | 26 |
|  | Severe |  | 8 | 2.4 |
| Grades | Satisfactory |  | 214 | 62.9 |
|  | Unsatisfactory |  | 126 | 37.6 |
| Teacher support | Good |  | 312 | 92.3 |
|  | Moderate |  | 21 | 6.2 |
|  | Poor |  | 5 | 1.5 |
| Family-environmental |  |  |  |  |
| Relationship with parents | Good |  | 263 | 77.4 |
|  | Moderate |  | 71 | 20.9 |
|  | Poor |  | 6 | 1.7 |
| Relationships between family members | Good |  | 257 | 75.6 |
|  | Moderate |  | 79 | 23.2 |
|  | Poor |  | 4 | 1.2 |
| Overprotective parents | Yes |  | 279 | 81.8 |
|  | Moderately protective |  | 57 | 16.7 |
|  | No |  | 5 | 1.5 |
| Domestic violence | Not at all |  | 219 | 64.2 |
|  | Moderate |  | 112 | 32.8 |
|  | Severe |  | 10 | 2.9 |
| Family history of suicide | Yes |  | 17 | 5 |
|  | No |  | 321 | 95 |
| Socio-environmental |  |  |  |  |
| Bullying | Yes |  | 285 | 84.3 |
|  | No |  | 53 | 15.6 |
| Psychological | DASS-21 score | Category | n | % |
| Stress | 0-14 | No stress | 307 | 90 |
|  | 15-18 | Mild | 24 | 7.1 |
|  | 19-25 | Moderate | 10 | 2.9 |
|  | 26-33 | Severe | 0 | 0 |
|  | 34+ | Extremely severe | 0 | 0 |
| Anxiety | 0-7 | No anxiety | 280 | 82.1 |
|  | 8-9 | Mild | 26 | 7.6 |
|  | 10-14 | Moderate | 26 | 7.6 |
|  | 15-20 | Severe | 9 | 2.6 |
|  | 20+ | Extremely severe | 0 | 0 |
| Depression | 0-9 | No Depression | 280 | 82.1 |
|  | 10-13 | Mild | 38 | 11.1 |
|  | 14-20 | Moderate | 22 | 6.5 |
|  | 21-27 | Severe | 1 | 0.3 |
|  | 28+ | Extremely severe | 0 | 0 |
| Suicidal behavior | Yes (SBQ-R 7 or more) |  | 42 | 12.3 |
|  | No (SBQ-R <7) |  | 290 | 87.7 |

### 3.2 Associated factors of suicidal behavior among adolescents

Sociodemographic, behavioral, psychological, institutional, family-environmental, and socio-environmental factors were used to identify the associated factors of suicidal ideation among adolescents. Age, relationship status, having step-parents, peer conflict, grades, teacher support, bullying, relationship with parents, the relationship between family members, overprotective parents, family history of suicide, stress, anxiety, and depression were found statistically significant with a p-value of <0.05 in chi-square test. These variables were further analyzed through a multivariate analysis using multiple logistic regression to adjust the confounding factors.

Relationship status, having step-parents, peer conflict, overprotective parents, family history of suicide, stress, and anxiety were found to be significantly associated with the development of suicidal behavior in preadults. Unmarried and single students were 20% less likely to generate suicidal tendencies. However, it was the highest among married adolescents (100%). Preadults having step-parents had a 6.5 times greater odds ratio than those who didn’t have. 62.5% of students who were experiencing severe peer conflict had suicidal tendencies which is higher than those who had moderate (14.8%) or no (9.9%) disputes with classmates. The overconcerned attitude of parents acted as a protective factor for the augmentation of suicidal attitudes among young adults. It was comparatively higher (40%) in students whose parents were negligent about their academic results than those having overprotective parents (9%). Family history of suicide also worked as a concerning factor for the suicidal behaviors of adolescents; a 2 times higher odds ratio was detected among those having at least one family member committed or attempted suicide. The percentage was commensurate with stress levels; 8.5% (26), 41.7% (10), and 60% (6) adolescents had suicidal behavior who were encountering normal, mild, and moderate stress respectively. An association of anxiety with suicidal behavior was also observed; 44.4% of adolescents suffering from severe anxiety revealed suicidal tendencies which is greater than in other classes. ***Table 2***.

**Table 2.**
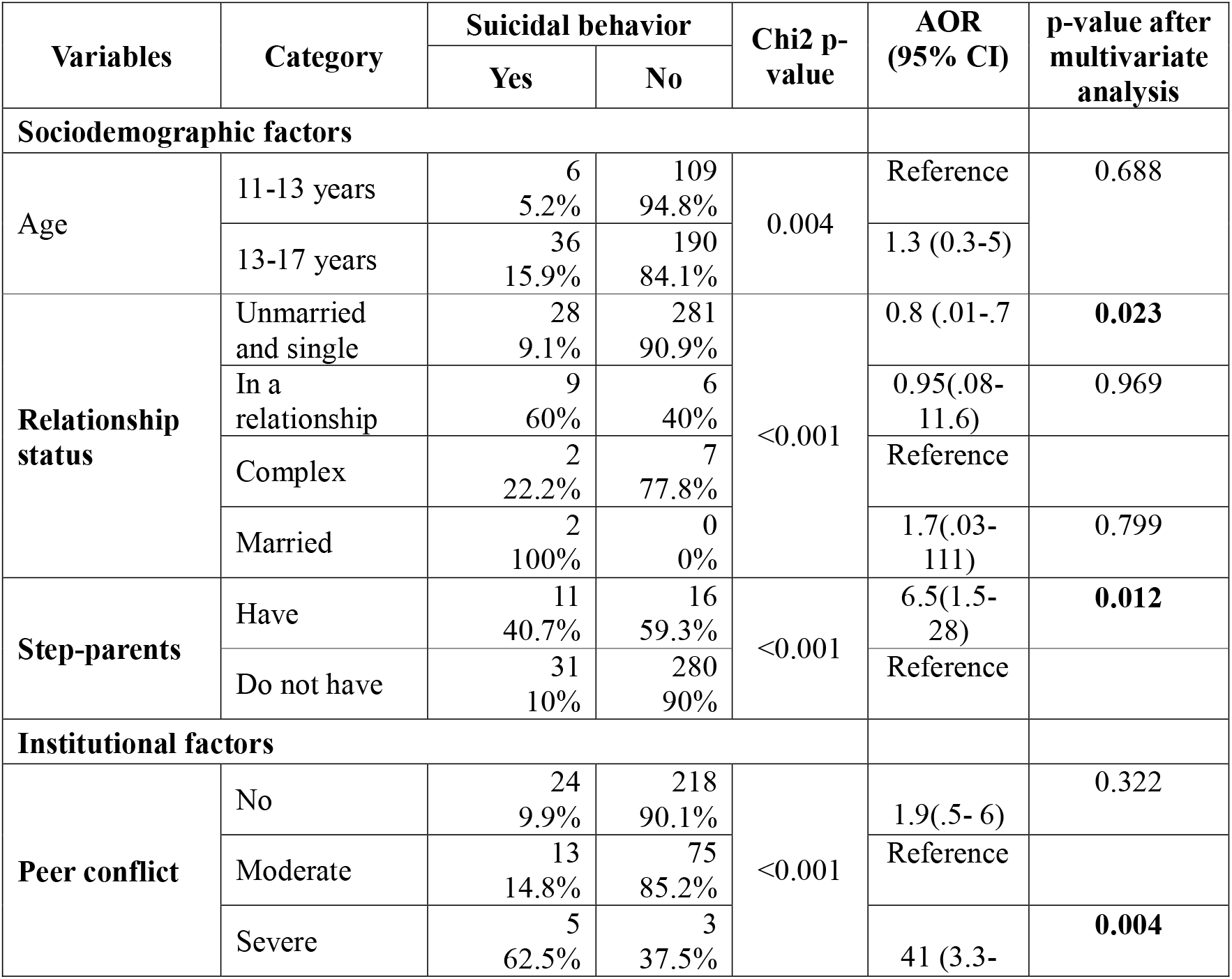
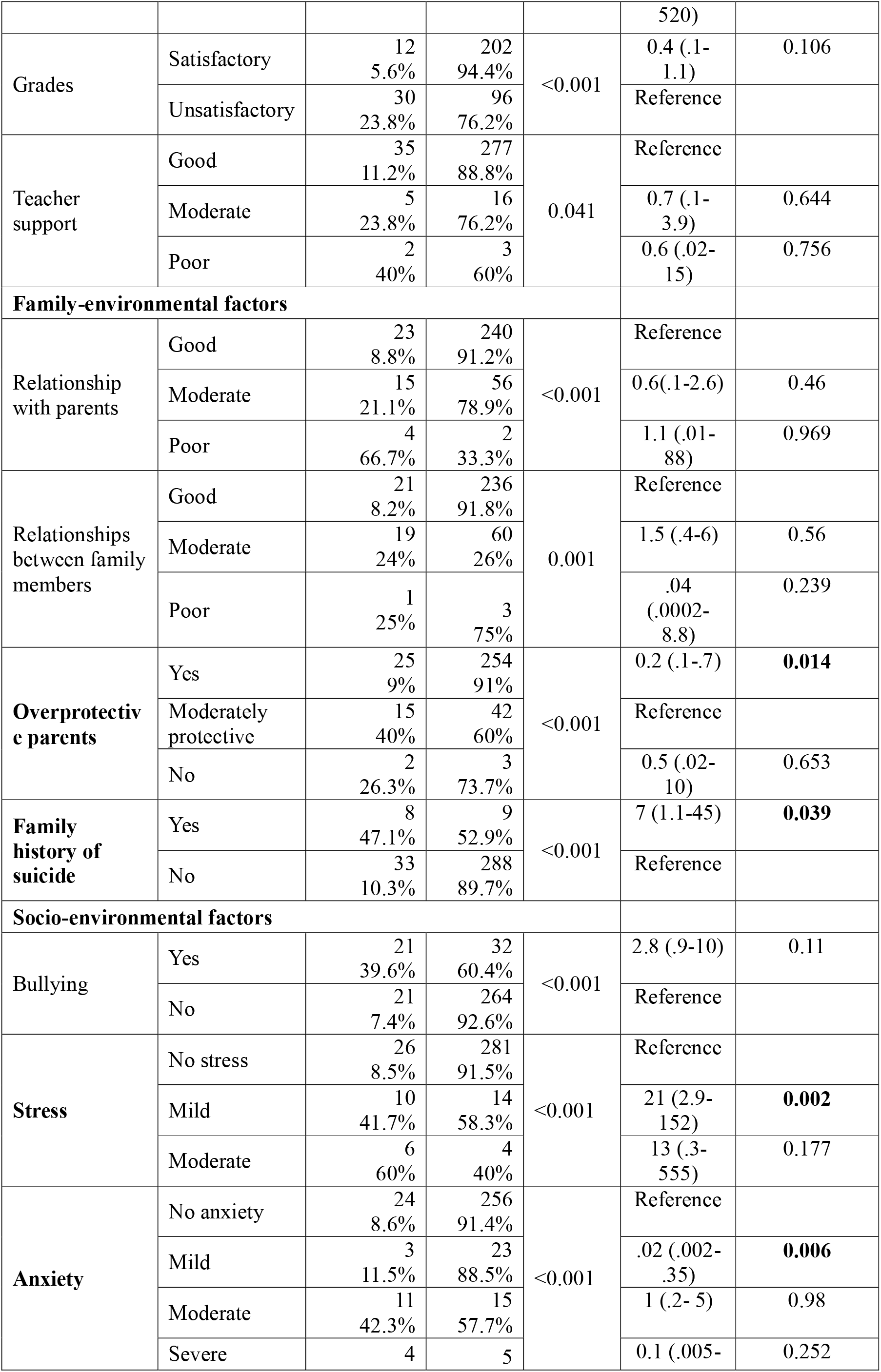
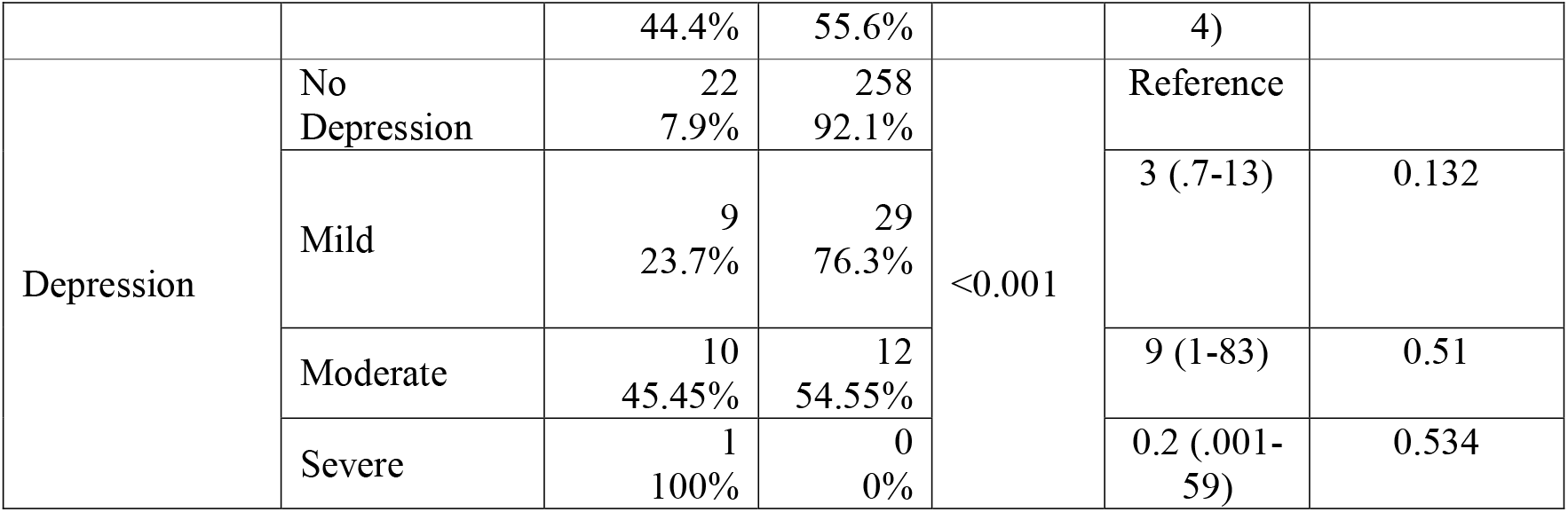
Factors associated with suicidal behavior among students of 11 to 17 years in Lohagara (a rural subdistrict in Bangladesh), 2024 (bivariate analysis by chi-squared test and multivariate analysis by multiple logistic regression).

## 4. Discussion

This study aimed to assess the prevalence and associated factors of developing suicidal behavior among adolescents in rural Bangladesh. The point prevalence of suicidal behavior in rural adolescents is detected as 12.3% (95% CI: 9.2% to 16.2%). This percentage is attributed to several significant factors including having step-parents, relationship status, stress, anxiety, peer conflict, and a family history of suicide. The overprotecting attitude of parents plays a pivotal role in nullifying the suicidal behaviors of their offspring.

The obtained 12.3% prevalence from this study aligns with the global prevalence (14%) of suicidal ideation among 12 to 17-year-olds (Biswas et al., 2020) and the prevalence in Brazilian adolescents (14.1%) (Souza et al., 2010). The evaluated result from this research is also in agreement with the research conducted by Khan et. al unveiling an 11.7% prevalence of suicidal behavior among 11 to 18-year-old adolescents in Bangladesh (Khan et al., 2020) and the research carried out by Tasnim et. al during the onset of Covid-19 that highlighted a 12.8% prevalence (Tasnim et al., 2020). However, the result of this study is somewhat distant from the study conducted by Begum et. al, which revealed 5% of suicidal ideation among 14 to 19-year-olds in rural Bangladesh (Begum et al., 2017), and that is probably due to the variation in time, methodology, and age groups.

A study conducted by Ang et. al depicts that adolescent boys living in single-parent families are more prone to develop suicidal ideation than their counterparts (Ang & Ooi, 2004). The current study found the history of having step-parents play a key role in evolving suicidal behavior in young adults. Married, promised-to-get-married, or those who have received marriage requests are more prone to generate suicidal thoughts than adolescent girls who haven’t undergone the marriage process (Gage, 2013). This research depicts a significant association between relationship status and suicidal behavior in young adults.

Lack of peer support has a decisive role in preadult suicidal thoughts development (Fotti et al., 2006; Khan et al., 2020). In a study, Biswas et. al highlighted peer conflict as a reinforcing factor for the development of suicidal behavior in adolescents (Biswas et al., 2020), which is in line with this study.

Stress(Tasnim et al., 2020) and anxiety(Khan et al., 2020; Tasnim et al., 2020) are important psychological contributors having a substantial role in growing suicidal thoughts in adolescents and this study is in agreement with this.

Parenting style plays a crucial role in deterring suicidal thoughts in offspring. Some studies illustrate that the overprotective attitude of parents helps minimize suicidal behaviors in young adults (Gau et al., 2008; Singh & Behmani, 2018). School-going-children, whose parents check their homework regularly have less likelihood of generating suicidal ideation (Cheng et al., 2009; Khan et al., 2020). This study is in line with this finding. On the other hand, the current study found a similarity with studies that determined the family history of suicide as a key factor in developing suicidal behavior in preadults (Cerel & Roberts, 2005; Wagner et al., 2003).

Some studies found significant associations between loneliness (Pandey et al., 2019), bullying (Peprah et al., 2023) depression, and gender (Rudatsikira et al., 2007) with the development of suicidal tendencies in adolescents. This study doesn’t find any considerable association with them, perhaps because of the distinction in age groups, locations, and sample sizes.

## 5. Limitation

This study was conducted with data collected from a specific school in an area because of the time and resource limitations. Though the school admission policy allows diversity, students from remote areas of Lohagara aren’t inclined to get enrolled here. For this reason, this study doesn’t fully capture the divergence of the community people and thus leaves a chance of generalizability bias.

## 6. Conclusion

However, about 1 in 10 students are developing suicidal behavior at an early age, which is a matter of concern. Further research with students from different locations in Bangladesh will help identify other associated or area-specific factors if available. Stakeholders and policy-makers should come forward to draft plans and policies to scale down the burden. In addition, ensuring the participation of young adults in health promotional and health educational programs organized on this issue and increasing their creative opportunities may keep their mental health well and safeguard them from emerging suicidal tendencies at school age.

## Data Availability

Data is available on figshare.com. DOI:10.6084/m9.figshare.25854445

https://figshare.com/articles/dataset/Prevalence_and_associated_factors_of_suicidal_behavior_of_adolescents_in_rural_Bangladesh/25854445

